# The DNA methylation Profile of Undifferentiated Arthritis Patients Anticipates their Subsequent Differentiation to Rheumatoid Arthritis

**DOI:** 10.1101/2020.12.23.20248764

**Authors:** Carlos de la Calle-Fabregat, Ellis Niemantsverdriet, Juan D. Cañete, Tianlu Li, Annette H. M. van der Helm-van Mil, Javier Rodríguez-Ubreva, Esteban Ballestar

**Affiliations:** Epigenetics and Immune Disease Group, Josep Carreras Research Institute (IJC), Barcelona, Spain; Department of Rheumatology, Leiden University Medical Center, P.O. Box 9600, 2300 RC Leiden, the Netherlands; Rheumatology Service, Hospital Clinic and Institut d’Investigacions Biomèdiques August Pi i Sunyer (IDIBAPS), 08036 Barcelona, Spain

**Author notes:** To whom correspondence should be addressed: E. Ballestar.

**Keywords:** undifferentiated arthritis, rheumatoid arthritis, DNA methylation, machine-learning

## Abstract

**OBJECTIVE:** Undifferentiated arthritis (UA) is the term used to cover all the cases of arthritis that do not fit a specific diagnosis. A significant percentage of UA patients progress to rheumatoid arthritis (RA), others to a different definite rheumatic disease, and the rest undergo spontaneous remission. Therapeutic intervention in patients with UA can delay or halt disease progression and its long-term consequences. It is therefore of inherent interest to identify those UA patients with a high probability of progressing to RA who would benefit from early appropriate therapy. We hypothesised that alterations in the DNA methylation profiles of immune cells may inform on the genetically- or environmentally-determined status of patients and potentially discriminate between disease subtypes.

**METHODS:** In this study, we performed DNA methylation profiling of a UA patient cohort, in which progression into RA occurs for a significant proportion of the patients.

**RESULTS:** We find differential DNA methylation in UA patients compared to healthy controls. Most importantly, our analysis identifies a DNA methylation signature characteristic of those UA cases that differentiate to RA. We demonstrate that the methylome of peripheral mononuclear cells can be used to anticipate the evolution of UA to RA, and that this methylome is associated with a number of inflammatory pathways and transcription factors. Finally, we design a machine-learning strategy for DNA methylation-based classification that predicts the differentiation of UA patients towards RA.

**CONCLUSION:** DNA methylation profiling provides a good predictor of UA-to-RA progression to anticipate targeted treatments and improve clinical management.

## INTRODUCTION

Undifferentiated arthritis (UA) is a common form of arthritis that involves joint inflammation but cannot be classified within any definite rheumatologic disorder. Eventually, around 30% of the UA patients will develop rheumatoid arthritis (RA) or other differentiated forms of arthritis, whereas 45-55% of the patients will achieve spontaneous remission [1]. UA represents a unique window of opportunity to intervene during the course of the disease before more severe manifestations become established.

The ability to provide early indicators for the treatment of UA patients with high risk of developing RA is of utmost relevance for the decision of timing and treatment type with disease modifying antirheumatic drugs (DMARDs), which usually hamper RA progression but are not recommended for UA patients who undergo eventual remission [2]. For that aim, prediction rules have been proven to be crucial tools to provide guidance to clinicians by estimating patient outcome probabilities. In fact, a prediction rule for UA patients has been previously developed, strictly based on patient clinical data [3]. This model accurately estimates the risk of developing RA in more than 75% of the patients with UA. However, clinical data-based rules, although easy to implement in the clinical setting, usually fail to provide a detailed biological basis for individual phenotypic presentations of the disease, and usually do not succeed in the totality of predictions. In this regard, approaches including omics data may provide compelling alternatives or complementary tools for both improving prediction accuracy and allowing an in-depth characterization of the molecular alterations of patients [4].

Epigenetic alterations are associated with both genetic and environmentally-driven determinants which can in turn characterize pathogenic phenotypes. Specifically, DNA methylation and histone modifications, that are altered in multiple pathological contexts, are proposed to be both a causal factor [5] and a consequence of disease [6], as well as an intermediary for genetic susceptibility [7]. In all cases, the exhaustive study of these alterations through high-throughput technologies allows a detailed description and identification of novel molecular pathways that undergo alterations in a pathogenic context. DNA methylation is one of the most stable and easily comparable epigenetic modification, and thus stands out as an ideal candidate for biomarker discovery [8].

In the present study, we characterized the DNA methylome of patients with UA in comparison to healthy controls. We also analysed those data using different patient classification criteria, which proved to have a pivotal effect on DNA methylation profiles. Besides, we obtained the DNA methylation profiles of UA patients with known diverging future phenotypes. Moreover, we analysed the profiles of patients with definite RA and compared them to those from UA. The identification and interpretation of these data stand out as a valuable resource to delve into the molecular alterations of UA patients. Finally, we propose the use of DNA methylation data as a candidate biomarker with ability to improve clinical prediction rules by integrating molecular insights and clinical knowledge for the prediction of patient outcome.

## METHODS

### Patient cohort

The detailed descriptive and clinical data of the patients included in the study are summarised in Supplementary Table 1. For this study, we determined the patient diagnosis following both the American College of Rheumatology (ACR) 1987 criteria [9] and the ACR/EULAR (European League Against Rheumatism) 2010 criteria [10]. As presented in the first section of Results (see text below and Supplementary Figure 1), the comparison of RA and UA patients by the two criteria (1987 vs. 2010) showed more robust DNA methylation profiles using the 1987 classification criteria, supporting its use as a benchmark in the context of the present study.

**Figure 1.**
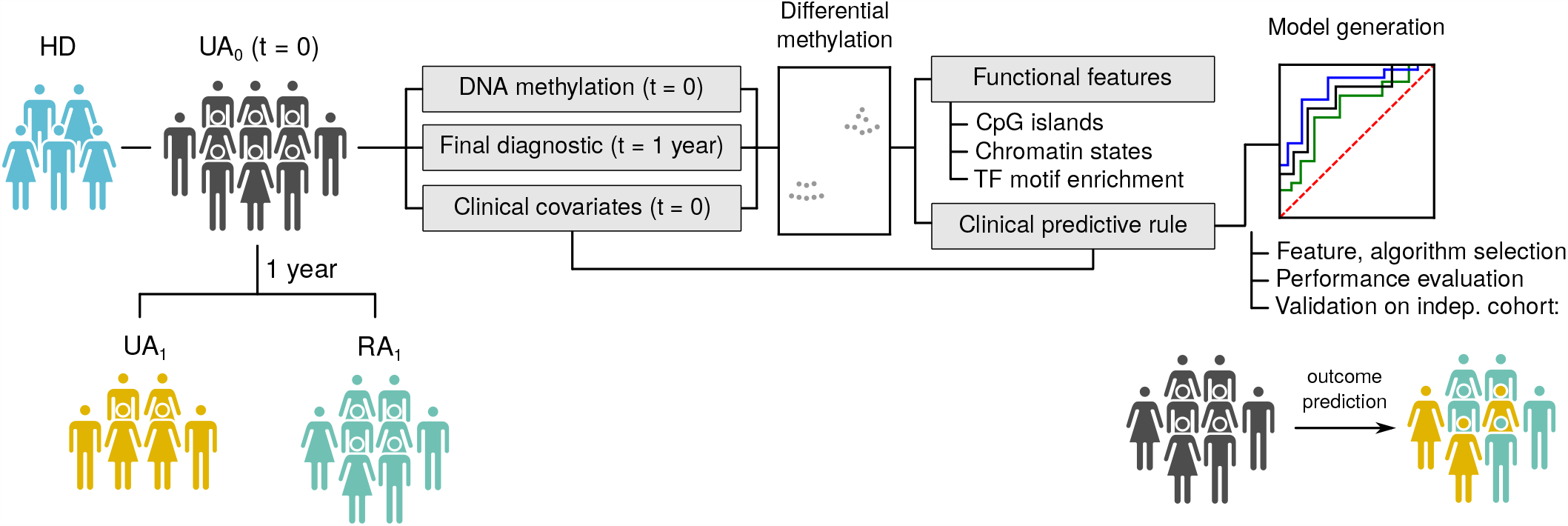
Study design. Conceptual and analytical workflow diagram. HD, healthy donor; UA_0_, undifferentiated arthritis at time = 0; UA_1_, undifferentiated arthritis at time = 1 year; RA_1_, rheumatoid arthritis at time = 1 year; TF, transcription factor.

### DNA Methylation profiling

Infinium HumanMethylation 450K and EPIC BeadChip arrays (Illumina) were used to analyse DNA methylation. These microarrays interrogate ∼450,000 and ∼850,000 CpG sites per sample, covering ∼96% of CpG islands and ∼99% of annotated RefSeq genes, distributed across distinct genic regions (5’UTR, TSS, gene body, 3’UTR and intergenic). Genomic DNA from peripheral blood mononuclear cells (PBMCs) was extracted and converted with sodium bisulfite using EZ DNA Methylation™kit (Zymo Research, Orange, CA). Bisulfite-converted DNA was hybridised on the arrays following manufacturer’s instructions, and fluorescence intensities were imaged using a BeadArray Reader (Illumina). Image processing, intensity data extraction and methylation value estimation procedures were performed as described in [11]. For downstream analysis and data visualization, M and beta (B) values were used, respectively. M values are calculated as the base 2 log ratio of the intensities of the methylated and unmethylated probes. B values are calculated as the ratio of the methylated probe intensity to the overall intensity (sum of the methylated plus the unmethylated probe intensities). For statistical purposes, the use of M values is recommended, since B value distributions display heteroscedasticity in low and high methylation ranges [12]. B values, which range from 0 (0% methylation) to 1 (100% methylation) are used for visualization purposes. The raw and normalized DNA methylation data reported in this paper is deposited in the Gene Expression Omnibus (GEO) database.

### Quality control, data normalization and statistical analysis of DMPs and DVPs

Methylation data were processed and normalized using the R statistical language. Preprocessing and quality control methods used in this study are contained in the R libraries *minfi* and *lumi* [13,14]. Significant probes were filtered by a detection p value (< 0.01) and normalization of methylation values was carried out with Illumina method. CpGs in single nucleotide polymorphism (SNP) loci were excluded from the analysis. To avoid discordant information among different samples, sex chromosomes (X and Y) were also excluded from the analysis. Since data was generated in a single batch, there was no need for a batch effect correction.

M values were used to identify significant differentially methylated CpGs between groups by empirical bayes-moderated t-test method, contained in the *limma* package [15]. FDR (false discovery rate) < 0.05 and p value < 0.05 where used as significance thresholds for the identification of DMPs in HD vs. UA_0_ and UA_1_ vs. RA_1_ comparisons, respectively. To overcome the confounding effect of age differences among sample groups, age-correlated CpGs in peripheral blood samples [16] were removed from all analysis derived from HD vs. UA_0_ comparison. Before all comparisons, cell type composition was computationally estimated in every sample with *estimateCellCounts* function from *minfi*, and sample groups were balanced by cell type composition average. Differentially methylated regions (DMRs) were identified using the *bumphunter* function from the *minfi* package, in which a FDR < 0.05 in regions containing 3 or more CpGs were considered DMRs. Differentially variable positions (DVPs) were identified by functions from the *iEVORA* package [17]. *iEVORA*’s algorithm identifies differential variance by applying a Bartlett’s test (q value < 0.001), followed by a mean comparison t-test (p value < 0.05), to regularise variability test, which is overly sensitive to single outliers.

### Gene Ontology, motif enrichment and chromatin state enrichment analysis

Gene ontology (GO) analysis was conducted with Genomic Regions Enrichment of Annotations Tool (GREAT) [18], in basal plus extensions mode with default settings, using array annotation as background regions. GO categories with FDR < 0.05 in the hypergeometric test were considered significantly enriched.

Transcription factor motif enrichment analysis (TFME) was performed using the HOMER software [19]. A flanking window of 250 bp was added to DMP coordinates, and array annotation was used as background. Uppermost enriched factors are depicted in the article figures.

Chromatin functional states enrichment analysis of DMPs was performed by using public PBMC data from Roadmap Epigenomics Project (http://www.roadmapepigenomics.org/) generated with ChromHMM [20]. We used a primary chromHMM dataset, containing 15 chromatin states based on 5 histone modification data. Enrichments were calculated with a Fisher’s test using array annotation as background regions. Only significantly enriched states are shown.

### Machine learning model construction and optimization

We developed binary outcome predictive models from DNA methylation and clinical data, by applying logistic regression, random forest and support vectors machine (SVM) algorithms, using functions from the *caret* R package [21]. For the internal validation of all models, data was randomly split in train and test subsets (representing ¾ and ⅓ of the original dataset, respectively). In order to reduce data dimensions, most significant DMPs in UA_1_ vs RA_1_ comparison, identified by *limma* in train dataset samples, were selected as predictors. Different predictive models were built using increasing amounts of CpGs as predictors (1 to 50 CpG, by 1) with or without the addition of clinical covariates, and probabilities of RA/UA diagnosis at year 1 were calculated for every patient within the test set. Highly correlated CpGs (Pearson’s correlation coefficient *r* > 0.5) were excluded, and a 10-fold cross-validation was performed on all models. Prediction on the test set was performed by selecting most frequent CpG predictors after cross-validation. The best models were selected by mean accuracy of prediction on the test set. Models were evaluated by comparing their area under the receiver operating characteristics (ROC) curve (AUC). Finally, for external validation, we analysed DNA methylation data of an independent cohort of 8 patients and RA/UA diagnosis at year = 1 was estimated with the previously selected models. For reproducibility purposes, before any analysis that required random sampling, the random number seed was set at a value of 12345 with the *set*.*seed* function.

### Heatmaps, PCA, *k*-means clustering and plots

Heatmaps of DMPs were generated with functions from the *ComplexHeatmap* and *gplots* R Packages. In all cases, blue and red indicate low and high normalized DNA methylation, respectively. PCA projection matrices were calculated with R’ s *prcomp* function and visual representations of PCs were plotted with *ggfortify* package functions. *k*-means clustering was performed with functions from the *factoextra* package. Selection of the optimal number of clusters was based on Gap clustering evaluation [22]. Heatmaps and PCA from DMP analyses that required the inclusion of covariates in the model (Figure 3C, Figure 5, Supplementary Figure 1 and Supplementary Figure 5; specified covariates at figure legends) were derived from B values corrected for the same covariates with the *removeBatchEffect* function from *limma* package. Box, bar, violin, bubble, scatter, line and ROC curve plots were generated using functions from the *ggplot2* and *ggpubr* packages, and depicted significance was calculated with R’ s *stat* packages.

## RESULTS

### Undifferentiated arthritis PBMCs display an altered DNA methylome in inflammation-related genes

Firstly, we obtained the DNA methylation profile of PBMCs from a cohort of UA patients (Figure 1). Descriptive and clinical variables of these patients, including age, sex, seropositivity for anti-citrullinated antibodies (CCP2), rheumatoid factor (RF) and disease activity (DAS44) were also collected (Supplementary Table 1). Since UA/RA patients can be classified according to two main criteria, ACR 1987 [9] and ACR/EULAR 2010 criteria [10], we decided to compare the DNA methylation profiles between patients’ diagnosis following both classifications. In the first place, we identified the most significant differences between patients that were classified either as UA or RA by both criteria (considered as *bona fide* UA or RA), named Group 1 and 3 respectively. After that, we performed principal component analysis (PCA) and jointly plotted the data of patients that undergo a change in group between criteria: patients diagnosed as UA in 1987 and as RA in 2010 were named Group 2 and patients diagnosed as RA in 1987 and as UA in 2010, Group 4 (Supplementary Figure 1). There was an almost total overlap between Groups 1 and 2, and between Groups 3 and 4, suggesting that the 1987 criteria identifies more homogeneous epigenetic signatures of patients than the 2010 criteria. For that matter, we deemed appropriate for our analyses to stratify patients following ACR 1987 diagnostic criteria.

In total, 64 UA patients (labelled as UA_0_) and 13 healthy donors (HD) were analysed (an illustrated flow chart of the study design is depicted in Figure 1). Following data correction for age and sample balancing by sex and cell type proportions (Methods and Supplementary Figure 2A), the comparison between UA_0_ and HD revealed 321 hypermethylated and 3029 hypomethylated CpG sites (Figure 2A). Gene ontology (GO) analysis of the differentially methylated positions (DMPs) revealed enrichment for multiple categories regarding inflammatory response, immune cell activation, vitamin metabolism, and cytokine and chemokine signalling pathways in both clusters (Figure 2B). Among those, interleukin (IL)-1, 6, 12, and 10, tumour necrosis factor (TNF), macrophage colony-stimulating factor (CSF-1) and chemokine (C-X-C motif) ligand 2 (CXC2L) signalling pathways were shown to be enriched within the affected regions. The hypomethylated regions were specifically enriched in categories related to antimicrobial response and IFN-type I production. Detailed examples of methylation of CpG sites proximal to genes contained in GO categories are depicted in Figure 2C. Two examples of differentially methylated regions (DMRs) are shown in Supplementary Figure 2B. These genes were selected by their direct previously reported involvement in rheumatic diseases and their underlying molecular pathways. For instance, we found differences in CpG sites located in cytokine and chemokine genes, such as *CXCR5, IL10, IL1R1* and *IRAK2*; TNF signalling pathway genes, such as *LTA, TNFSF10* and *TRAF4*; interferon-type I-activated transcription factor IRF8, and others.

**Figure 2.**
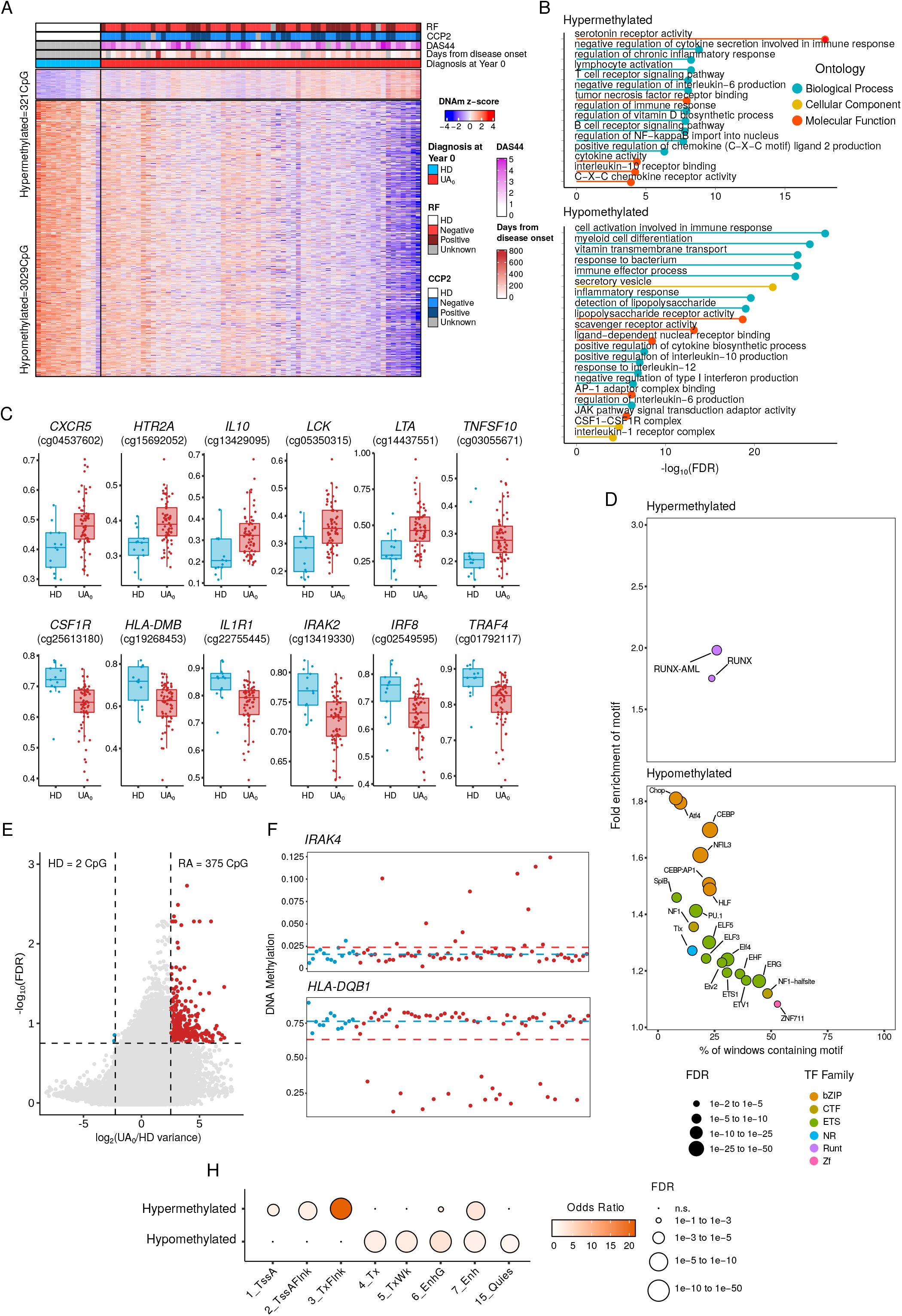
Characterization of the DNA methylation profiles of UA_0_ patients compared to HD. (A) Heatmap showing DMPs between UA_0_ and HD (FDR < 0.05). Clinical covariates of each patient are shown at the top of the heatmap. A legend scale of normalized DNA methylation is shown at the right, with red indicating lower methylation and blue indicating higher methylation. Legend scales are shown for all included clinical covariates at the right of the heatmap. (B) Significant GO categories selected from the analysis with GREAT of the identified DMPs. (C) B values of selected CpG sites contained in the displayed GO categories. (D) Significantly enriched motifs in DMPs from both clusters, analysed with HOMER. (E) Variability plot depicting log_2_ratio of variance (var_UA0_/var_HD_) for individual CpG base 10 log FDR of the mean comparison t-test. Significant DVPs for both groups identified by the *iEVORA* package are highlighted in color. (F) Two selected examples of DVPs, displaying individual DNA methylation. (G) Chromatin functional state enrichment based on public PBMC data from Roadmap Epigenomics Project. RF, rheumatoid factor; CCP2, anti-citric citrullinated peptide antibody; DAS44, disease activity score 44; HD, healthy donor; UA_0_, undifferentiated arthritis at year 0; FDR, false discovery rate; TF, transcription factor; bZIP, basic leucine zipper domain family; NR, nuclear receptor family; Zf, zinc finger domain family; CTF, CCAAT box-binding transcription factor family.

**Figure 3.**
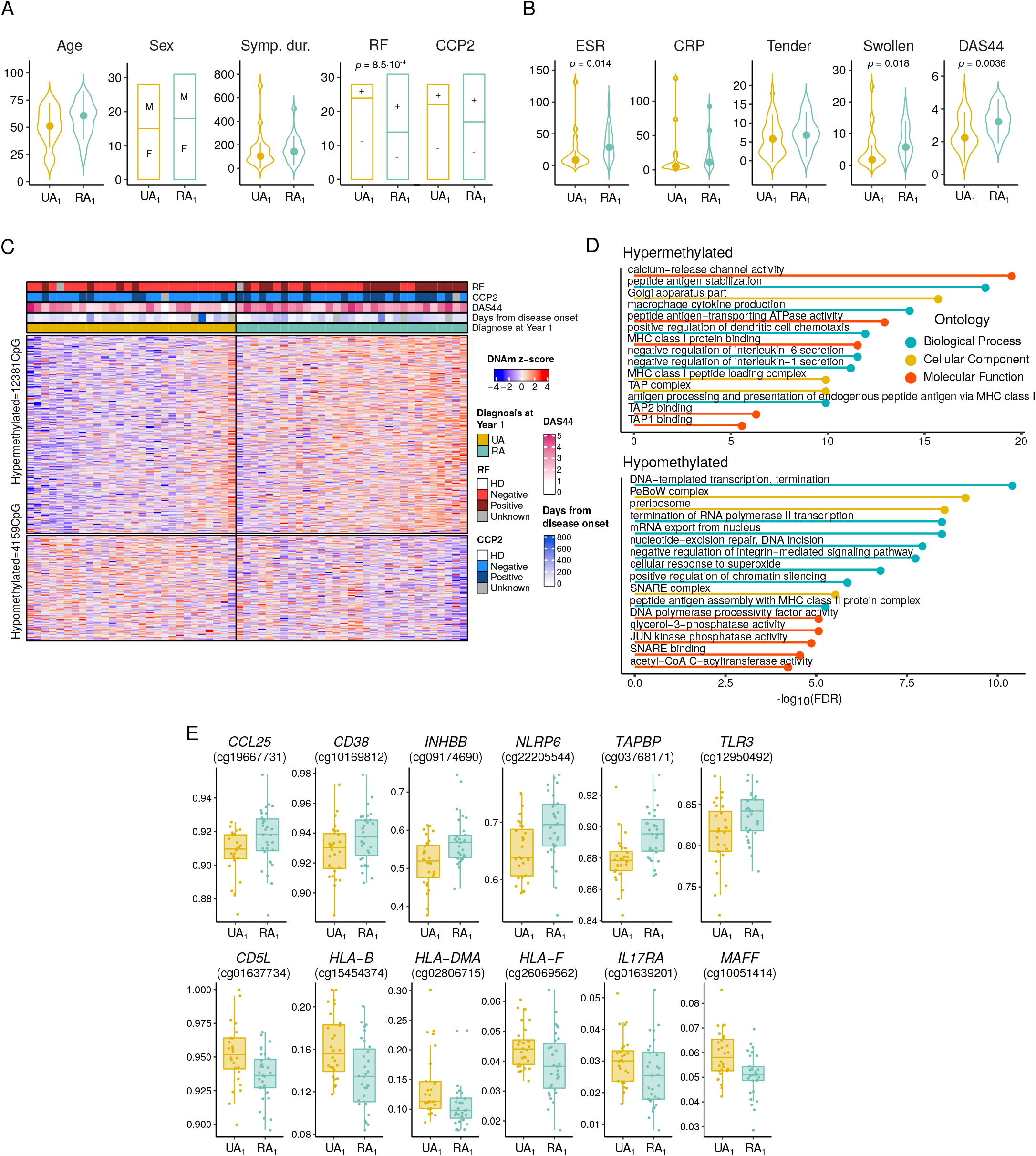
Characterization of the DNA methylation profiles of patients with diverging outcomes after 1 year. (A) Descriptive and clinical variables of UA_1_ and RA_1_ groups. (B) Clinical variables of UA_1_ and RA_1_ included in the DAS44 formula. Statistical significance in (A) and (B) is derived by Wilcoxon tests for numeric variables and by chi-squared tests for categorical variables. (C) Heatmap showing DMPs between UA_1_ and RA_1_ (p value < 0.05). Clinical covariates of each patient are shown at the top of the heatmap. A legend scale of normalized DNA methylation is shown at the right, with red indicating lower methylation and blue indicating higher methylation. Legend scales are shown for all included clinical covariates at the right of the heatmap. (D) Significant GO categories selected from the analysis with GREAT of the identified DMPs. (E) B values of selected CpG sites contained in the displayed GO categories. Symp. dur, Symptoms duration in days; RF, rheumatoid factor; CCP2, anti-cyclic citrullinated peptide antibody; ESR, erythrocyte sedimentation rate; CRP, C-reactive protein; DAS44, disease activity score 44; HD, healthy donor; UA_1_, undifferentiated arthritis at year 1; RA_1_, rheumatoid arthritis at year 1; FDR, false discovery rate.

Analysis of transcription factor (TF) binding motifs revealed enrichment of motifs belonging to the RUNX and IRF TF families in the hypermethylated cluster. Within the hypomethylated cluster, motifs of TFs in bZIP and ETS families were predominantly enriched (Figure 2D).

Additionally, we performed a differentially variable position (DVP) analysis, which revealed a greater heterogeneity of DNA methylation within the UA_0_ group (Figure 2E). Those DVPs exhibited less than 2% overlap with the previously identified DMPs, further suggesting the presence of intrinsic variance within the UA_0_ group (Supplementary Figure 2C). Figure 2F depicts examples of two DVPs proximal to genes related to the previously identified GO categories. These data suggest the existence of an underlying epigenetic heterogeneity of UA patients, which might play a role in the diverse clinical presentation of the disease in each individual.

To ascertain the relationship between DNA methylation and genomic function, we calculated enrichment of the identified DMPs in 15 distinct chromatin states, defined by combinations of epigenetic modifications in PBMCs [23] (Figure 2H). DMPs in the hypermethylated cluster were enriched in regions containing gene coding sequences (CDS) and transcription start sites (TSS), while hypomethylated DMPs are enriched in actively transcribed regions. Both clusters display an enrichment in enhancer regulatory regions, in keeping with previously published studies focused on the analysis of dynamic DNA methylation [24].

### The methylome of UA patients anticipates subsequent evolution of the disease

The higher variability of the methylation profiles of UA_0_ samples in comparison with healthy individuals is in agreement with the clinical heterogeneity of this group. UA_0_is composed of two subgroups, one with patients that undergo subsequent differentiation to RA (designated as RA_1_) and patients that remain undifferentiated (designated as UA_1_) one year after from the initial visit (Figure 1). In fact, slight clinical dissimilarities can be found among these two groups. For instance, RA_1_patients have a higher frequency of seropositivity for rheumatoid factor (RF) (Figure 3A). Besides, the disease activity score (measured by DAS44, [25]) and some of the parameters included in its calculation, of note, the ESR (erythrocyte sedimentation rate) and the number of swollen joints, are also higher in RA_1_ (Figure 3B). However, technically, such differences do not allow the identification of those patients as RA in the clinical setting. For that, we aimed at identifying DNA methylation alterations that might help to the anticipation of a future diagnostic in a prospective manner. In our analysis, we included the clinical features with significant differences among the two groups (RF and DAS44) as covariates, in order to identify methylation changes that were not due to the effect of those differences.

After cell type composition similarity verification (Supplementary Figure 3A), the comparison of the previous DNA methylation profiles in relation to the clinical groups defined one year later (UA_1_ vs. RA_1_) led to the identification of 12,381 hypermethylated CpGs and 4,159 hypomethylated CpGs (Figure 3C). DMPs in the hypermethylated cluster were enriched in GO categories related to antigen presentation through major histocompatibility complex (MHC) class I, as well as inflammatory cytokine signalling (such as IL-1 and IL-6). GO categories in the hypomethylated cluster were mainly related to basic cellular processes such as gene transcription, translation and metabolism, and an additional category related to antigenic presentation through MHC class II (Figure 3D). CpGs proximal to genes contained in the aforementioned GO categories and that are related to inflammatory cytokines and chemokine pathways, such as *CCL25, CD5L* and *IL17RA*, were selected and depicted in Figure 3E.

Analysis of TF binding motifs in the hypermethylated cluster revealed an enrichment of motifs belonging to BHLH and Zf families. The hypomethylated cluster showed enrichments of TFs from ETS and Zf families (Supplementary Figure 3B). Chromatin states enrichment for the hypermethylated cluster revealed an enrichment of regions located in active and poised TSS or their flaking regions. The hypomethylated cluster showed enrichment in either actively transcribed regions, enhancers and repressed chromatin (Supplementary Figure 3C). None of the chromatin states were commonly enriched in the two clusters, suggesting the involvement of distinct pathways underlying the identified alterations.

Taken together, these results describe for the first time a pre-established epigenetic signature of UA patients who will evolve to RA within 1 year.

### Incorporation of DNA methylation data improves the patient classification based on clinical parameters

Given our findings on the occurrence of DNA methylation differences between UA patient groups which had divergent diagnoses 1 year later, we investigated the possibility of using DNA methylation data to obtain predictive markers of disease progression. To this end, we applied machine learning approaches to build a classification system based on DNA methylation data or with the addition of clinical data. The pipeline of the methodology included a random split of the original data in ‘train’ and ‘test’ sets, followed by a selection of predictor CpGs, and a cross-validation for the internal evaluation of the model (Figure 4A). Models developed and evaluated through this procedure were constructed using logistic regression, random forest and support vectors machine (SVM) algorithms. Aiming at the selection of a relatively simple classifier, we generated models with increasing numbers of CpG sites as predictors (from 1 to 50 CpGs). In parallel, clinical data of patients (RF and DAS44) were included as explanatory variables. These variables, that showed significant differences among groups, were also included in previous studies describing classification rules that were strictly based on clinical information [3]. The comparison of the accuracies of all models (see Methods) evidenced the highest precision for SVM-generated models, with RF and DAS44 covariates included (Figure 4B). The top 10 most frequent CpGs (after performing 100-fold cross-validation) in the SVM+covariates model are shown in Figure 4C. Given that SVM+covariates models discriminate relatively well above 7 CpGs, we selected two examples of models representing one complex classifier, with 25 CpGs, and a simpler classifier, with 10 CpGs, that could potentially be implemented in the clinical setting. Finally, these models were applied to an external validation cohort (n = 8), recruited independently. The predicted outcome was compared to the observed outcome after 1 year for every patient (Figure 4D). Model with 10 CpGs was not able to increase the accuracy of the prediction by the clinical covariates alone (AUC = 0.625), but improved the accuracy of predicting the RA_1_ class. Model with 25 CpG+covariates predicted accurately the class of 7/8 patients (AUC = 0.875) (Figure 4D and Supplementary Figure 4A). In fact, models with > 25 CpGs predicted future diagnosis with average accuracy above 75%, and the addition of the CpG methylation data improved the predictive ability of clinical parameters in the majority of the models (Supplementary Figure 4B). Taken together, these results highlight the potential of adding DNA methylation as a diagnostic predictive biomarker.

**Figure 4.**
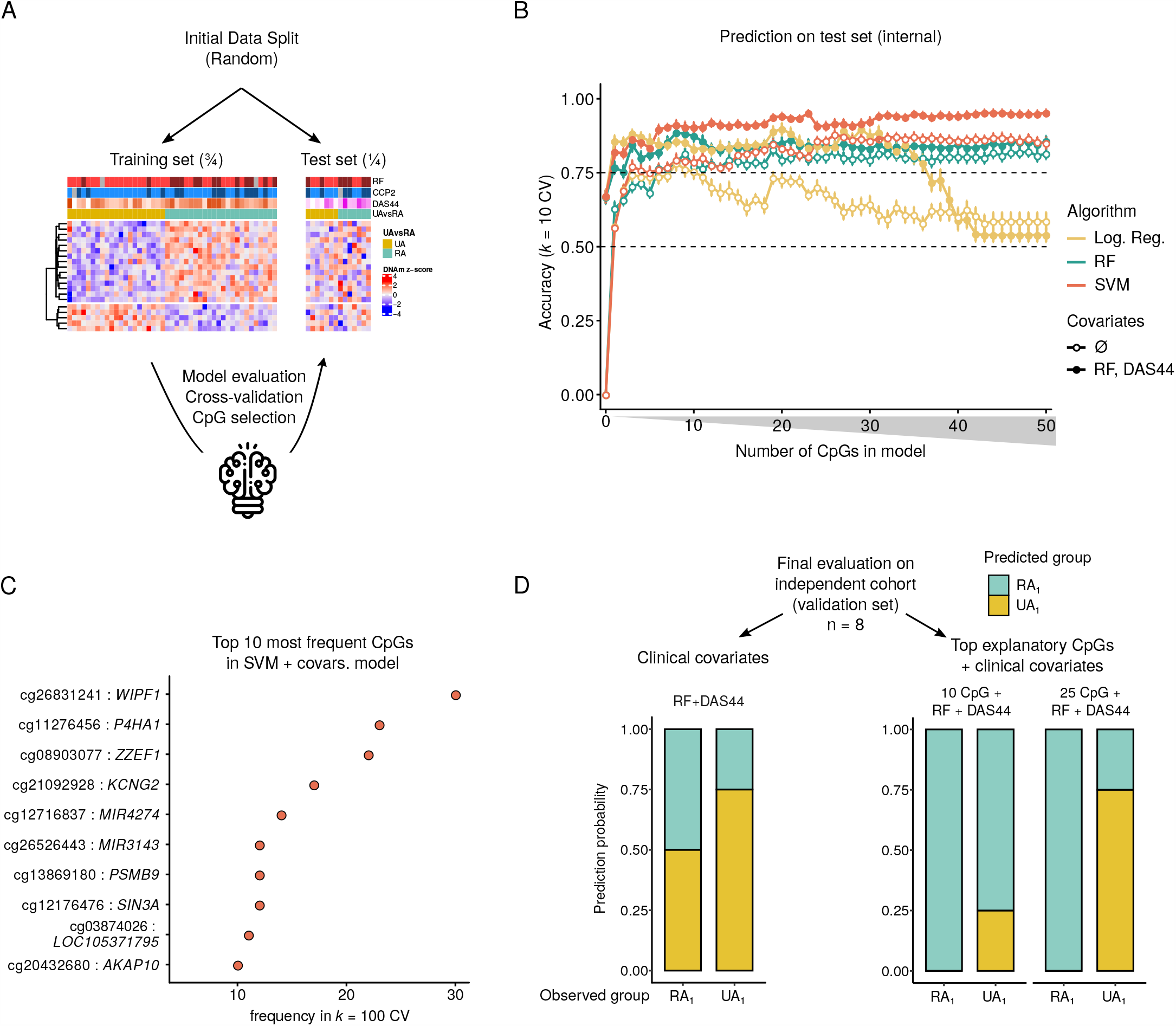
DNA methylation-based prediction rule by machine learning. (A) Schematic representation of the methodology, including feature (CpG) selection, evaluation of the model parameters and cross-validation. (B) Accuracy of the tested models across sequential numbers of most frequent CpGs as explanatory variables (1 to 50). For every CpG number, model accuracy mean and standard error from 10 independent cross-validations are shown. (C) Top most frequent CpGs after 100-fold cross-validation of the SVM model with covariates. (D) Classification results of selected models on an independent validation cohort. Barplot x-axis text indicates observed group, color indicates predicted group. Left, clinical covariates only-based prediction; right, DNA methylation plus clinical covariates-based models predicton. Log. Reg, logistic regression; RF, random forest; SVM, support vectors machine; RF, rheumatoid factor, DAS44, disease activity score 44.

**Figure 5.**
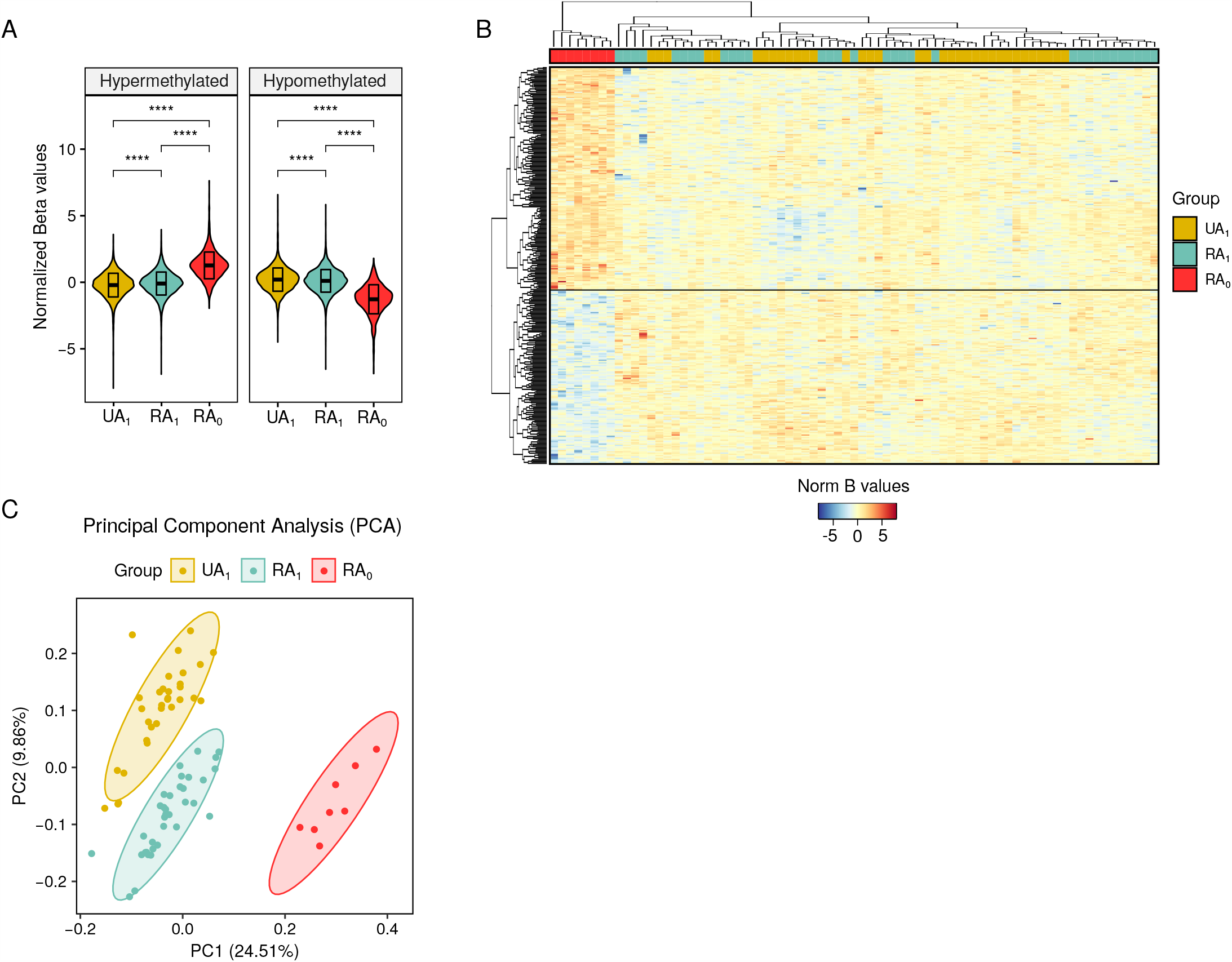
Comparison of UA_1_, RA_1_ and RA_0_ methylome profiles. (A) Violin plots showing normalized B value distributions of DMPs between UA_1_, RA_1_and RA_0_ profiles. Microarray model (450k or EPIC) was included as a covariate in the *limma* model. (B) Heatmap of the identified DMPs. Columns (samples) are clustered by a complete-linkage clustering algorithm. (C) Principal component analysis of the DMPs. Ellipsis show 95% confidence interval of the distribution of every group. UA_1_, undifferentiated arthritis at year 1; RA_1_, rheumatoid arthritis at year 1; RA_0_, rheumatoid arthritis at year 0.

### Comparison of UA with *bona fide* RA profiles reveals that RA_1_ patients are actually *en route* to RA status

To further characterize the UA_0_ subgroups, we compared the DNA methylation profiles of UA_0_ with those of terminally differentiated RA patients (diagnosed as RA at the time of visit, t = 0), labelled RA_0_. RA_0_ displayed the most distinct methylation profile, as shown by the greatest differences in mean DNA methylation when compared to both UA_1_ and RA_1_ (Figure 5A). We then performed unsupervised clustering of the significant DMPs among the three groups (RA_0_, UA_1_, RA_1_). We observed that all UA_1_ and RA_1_ (UA_0_) samples aggregate together in the same cluster, while all RA_0_ samples cluster independently (Figure 5B). Overall methylation of the identified DMPs showed significant differences among the three groups. In addition, these regions appear to experience a progressive dynamic from UA_1_ to RA_1_ to RA_0_, both for the hypermethylated and the hypomethylated clusters (Figure 5A). This tendency is further reinforced after reducing dimensionality of the DMP data by PCA, where RA_1_ patients lie in between RA_0_and UA_1_, which appear as the most extreme groups when projected in PC1 and PC2 (Figure 5C). Additionally, unsupervised *k*-means clustering of the DMP data reveals a total association of RA_0_ in an isolated cluster (Cluster 3), while UA_1_and RA_1_, which are largely associated to independent clusters (Clusters 1 and 2), show a certain degree of interspersing (Supplementary Figure 5A). After assigning samples to clusters, 5/32 (15.6%) of the UA_1_ samples belong to Cluster 2 (‘RA_1_’ cluster) while 7/35 (20%) of the RA_1_samples belong to Cluster 1 (‘UA_1_’ cluster). 8/8 (100%) of RA_0_ samples belong to Cluster 3 (Supplementary Figure 5B).

All these data reinforce the notion of a pre-existing RA-like epigenetic profile underlying some UA patients, which reveals a progression of these patients to RA status.

## DISCUSSION

Our analysis of the DNA methylomes of UA patients shows a distinct signature in comparison with healthy individuals, as well as specific differences between patients that subsequently evolve to RA and those that remain undifferentiated. These observations prompted us to design a machine learning-based method to predict the outcome of UA patients, which improves previous classification systems [3]. The finding that UA patients that will progress to RA have a more similar DNA methylation signature to well established RA patients supports the notion of pre-existing epigenetic signatures that might be used to anticipate the patient outcome and, therefore, improve therapeutic decisions.

In our cohort, the definition of UA is based upon having clinical arthritis but not meeting the specified classification criteria for RA. Therefore, the RA classification criteria used is critical to the interpretation of the findings. Before starting any analysis, classification criteria by ACR 1987 was compared to ACR/EULAR 2010 with regards to patient assignment to groups (UA/RA). Surprisingly, results from this analysis showed that DNA methylation profiles of patients cluster largely by the diagnostic provided by 1987 criteria. From this result, we presume that 1987 criteria might better reflect the dysregulated molecular status of the patients. Actually, some authors continue using this previous classification instead of the 2010 classification (which original purpose was to allow earlier diagnosis and treatment), which is less restrictive at including cases as RA and displays a low sensitivity at detecting ACPA negative RA cases when compared with 1987 or expert opinion [26,27].

Our results show for the first time that UA patients display epigenetic alterations when compared to healthy individuals. Those alterations, which occur in regions that are functionally associated with inflammatory pathways, are common to those previously observed in other inflammatory diseases. In particular, enriched functional categories of inflammation, immune cell activation and cytokine signalling are also found in RA [6,28,29], SLE [30,31], asthma [32] and IBD [33] in comparable studies, supporting the idea that UA shares epigenetic similarities with other inflammatory diseases and thus can be molecularly considered as such. Furthermore, the identification of vast DNA methylation differences at the TNF locus as well as alterations at several CpGs within the HLA class II region (both at the DMP and DVP level) reasserts UA as an arthritide, such as RA, with which shares clinical characteristics [7,34–36]

Furthermore, we identified DNA methylation differences among UA patients based on their prospective status, namely, the diagnosis after one year of the first visit (evolution to RA or persistent UA). After correcting by clinical features among the two groups, we were able to identify DNA methylation differences mainly localized in regions nearby genes related to inflammation and antigen presentation [37–40]. For instance, human leukocyte antigen (HLA) genes have been recursively linked to autoimmunity, showing both association with genetic susceptibility and epigenetic alterations in several studies [7,41,42]. HLA-B, HLA-DMA and HLA-F have further been linked to autoimmunity [43–45]. Other genes, such as CD38, have been shown to be upregulated at the protein level in UA patients and has been proposed as a therapeutic target of UA and early arthritis patients [46]. These observations suggest that those patients possess early biological alterations before undergoing diverging clinical fates. Disease onset identification in the clinical setting is often preceded by the presence of unapparent molecular triggers, as previously described for RA treatment response [47] and flare outbreaks [48]. Those determinants appear early in the disease course and cannot easily be detected by clinicians through non-invasive means. However, their sustained presence and effect at several levels may contribute to a specific pathological phenotype. We believe that this study underpins the potential of using epigenetic modifications as a molecular sensor for those early disease determinants in UA, in order to improve the classification criteria of UA patients and prevent damage caused by sustained inflammation.

Autoimmune arthritides are characterized by a high level of heterogeneity in terms of patient prognosis, joint damage, and response to treatment, for which mechanistic causality remains largely unknown [49]. In this sense, the use of high-throughput technologies has enabled the development of computational methods for the processing of patient omic data in search of novel and more precise conclusions [50–52]. For instance, the implementation of machine learning algorithms in high-dimensional data analysis has previously been used to improve stratification of patients [53–55] or to predict disease activity [56,57] in RA and SLE. In our study, we have used DNA methylation in addition to clinical data of UA patients by applying machine learning approaches, fine-tuning the prediction performance of previously existing classifiers [3] in an independent validation cohort. Our conclusions highlight the convenience of using both clinical and basic research data in conjunction for a complete and robust patient prognostic and therapeutic assessment. The results obtained herein are presented as a proof of concept to be further confirmed in independent studies with larger sample sizes. We hope our methodology can also be applied to other disease contexts.

The comparison of the methylation profiles of all the UA patients in our cohort (regardless their prospective status at 1 year, i.e. UA or RA) versus those initially classified as RA patients, showed that UA and RA individuals displayed differential methylation profiles, further supporting the idea that these two groups actually belong to distinct disease entities from a molecular/epigenetic perspective. Upon exploration by unsupervised analyses, both UA subgroups, UA_1_ and RA_1_, showed a higher resemblance compared to RA_0_, suggesting that despite the existing differences among them, both groups still behave as an entity when compared to a differentiated group. Interestingly, UA_1_ and RA_0_ were the most extreme groups, while RA_1_displayed an intermediate distribution. In all, these data suggest the pre-existence of a molecular/epigenetic signature in UA patients that undergo RA in the future.

Many efforts have been devoted to promptly abort the inflammatory process and the subsequent tissue damage in arthritic patients. Indeed, a delayed treatment of these patients is commonly associated with a worse global response to the treatments, joint destruction and impaired quality of life. In this regard, one of the benefits of the 2010 classification criteria settles in anticipating the progression of the disease to a more severe form, facilitating a rapid halt of the dysregulated inflammatory process and avoiding inflammation-associated tissue damage. However, this conservative approach might eventually constitute a double-edged sword since a premature treatment with DMARDs or other anti-inflammatory drugs, especially in patients that ultimately will not evolve to a more severe form of the disease, might be associated with undesired iatrogenic effects. In this context, our results regarding epigenetic signatures associated with distinctive UA evolution suggest that, in addition to specific clinical parameters, molecular features such as DNA methylation should be considered to be integrated into the clinics with the aim of a better classification of these patients.

## Data Availability

NA
We will upload the data to the NCBI GEO dataserver and have it opened before the paper is published

## Acknowledgements

The authors thank all the patients who graciously donated their time and samples to further rheumatoid arthritis research.

## Author contributions

C.C.-F., A.H.M., J.R.-U. and E.B. conceived experiments; C.C.-F. and J.R.-U. performed experiments; C.C.-F., and J.R.-U. performed biocomputing analysis; C.C.-F. and J.R.U. performed statistical analysis; E.N. and A.H.M. performed patient selection, provided samples and analysed clinical data; C.C.-F., E.N., T.L., J.C., J.R.-U., A.H.M. and E.B. analysed the data; C.C.-F., J.R.-U., and E.B. wrote the paper. All authors read and approved the final manuscript.

## Funding

We thank CERCA Programme/Generalitat de Catalunya and the Josep Carreras Foundation for institutional support. E.B. was funded by the Spanish Ministry of Science and Innovation (grant number SAF2017-88086-R), cofunded by FEDER funds/European Regional Development Fund (ERDF) - a way to build Europe. J.D.C was funded by a FIS grant (PI17/00993) from Institute of Health Carlos III (ISCIII). J.D.C. and E.B are supported by RETICS network grant from ISCIII (RIER, RD16/0012/0013).

## Competing interests

The authors declare no competing financial interests.

## Patient consent for publication

Not required.

## Ethics approval

The study was approved by the medical ethical testing committee (METC) Leiden Den Haag Delft (LDD), with cohort METC number: P11.210, and the board of the Bellvitge Hospital Ethical Committee (PR275/17).

## Data sharing statement

Methylation array data for this publication have been deposited in NCBI’s Gene Expression Omnibus and is accessible through GEO Series accession number.

## Notes

### Competing Interest Statement

The authors have declared no competing interest.

