## Supplementary figures and images for "The DNA methylation Profile of Undifferentiated Arthritis Patients Anticipates their Subsequent Differentiation to Rheumatoid Arthritis"

### Supp. Figure 1

Supplementary Figure 1

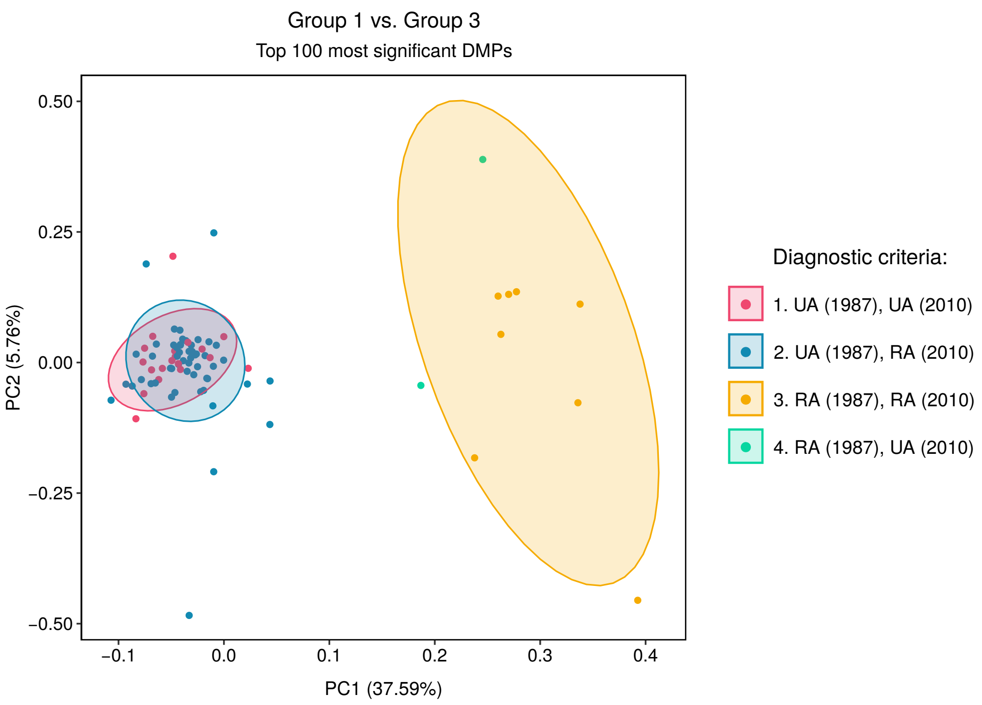

### Supp. Figure 2

Supplementary Figure 2

A

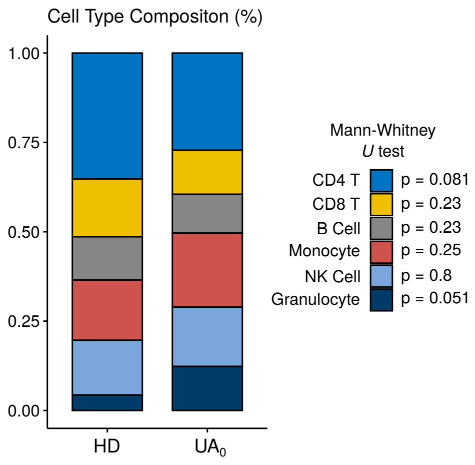


C

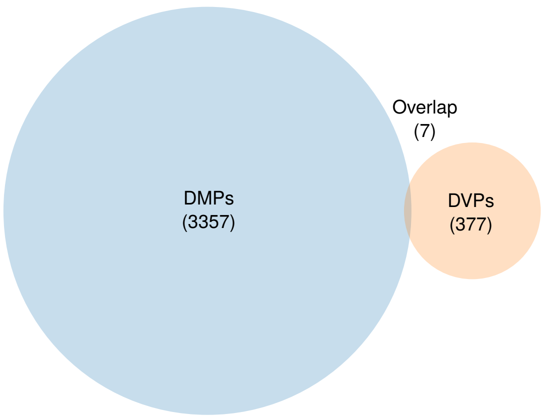


B

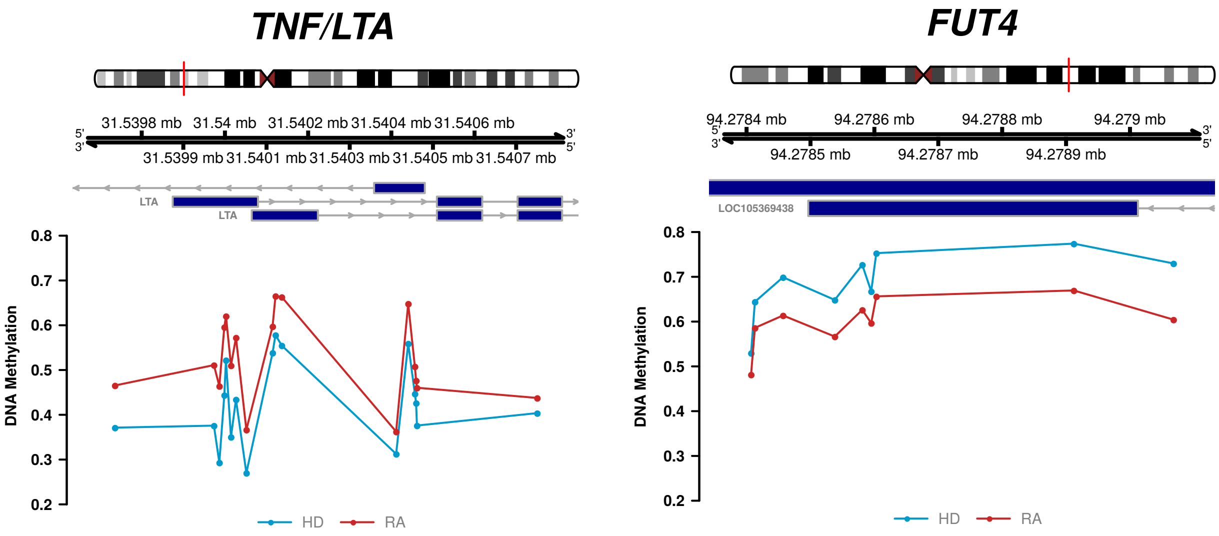

### Supp. Figure 3

Supplementary Figure 3

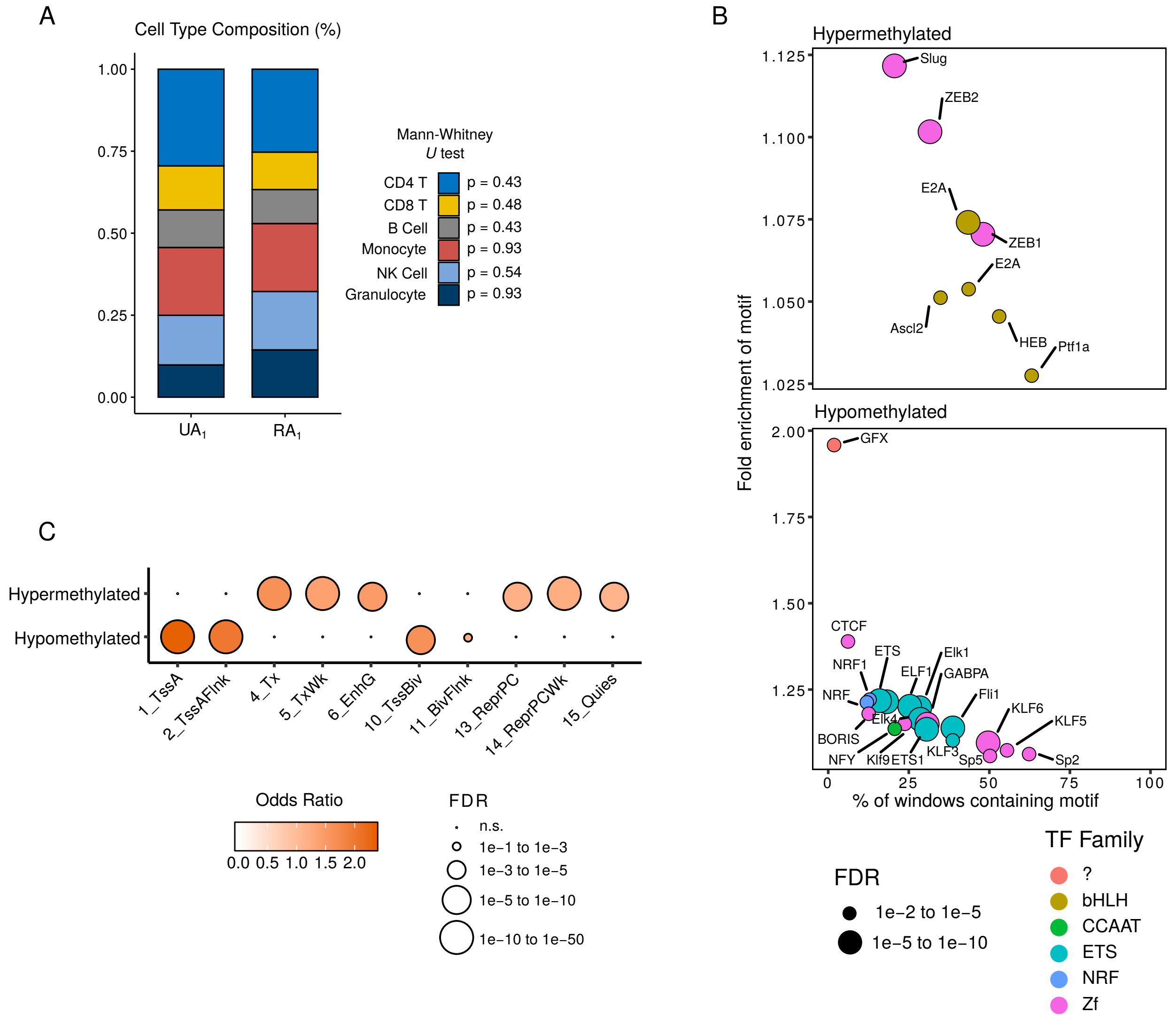

### Supp. Figure 4

Supplementary Figure 4

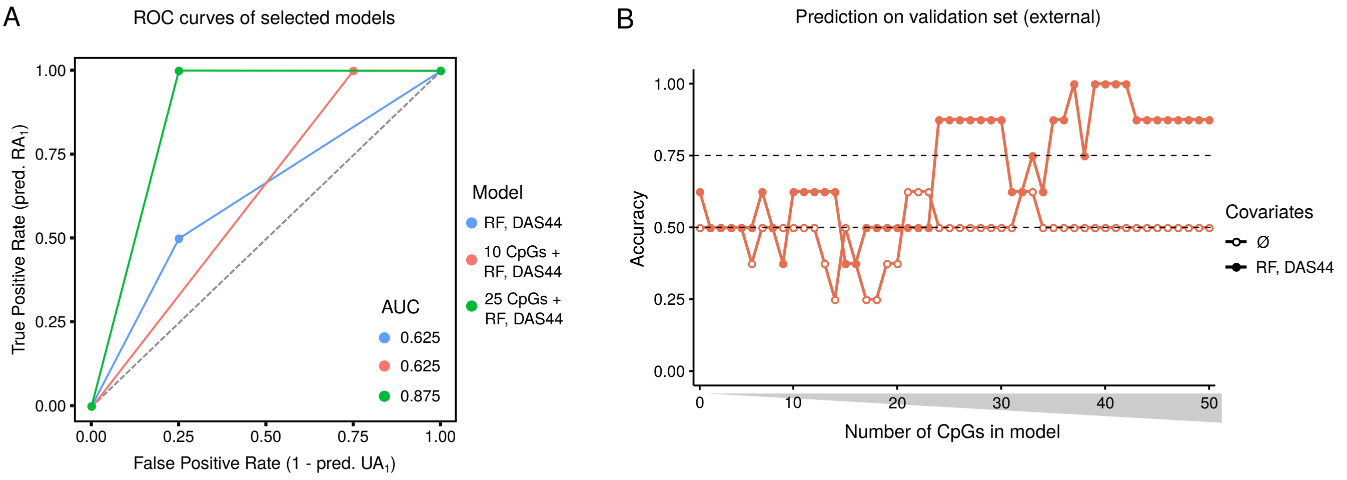
