## Supplementary material for "The DNA methylation Profile of Undifferentiated Arthritis Patients Anticipates their Subsequent Differentiation to Rheumatoid Arthritis": Supp. Figure 5

Supplementary Figure 5

A

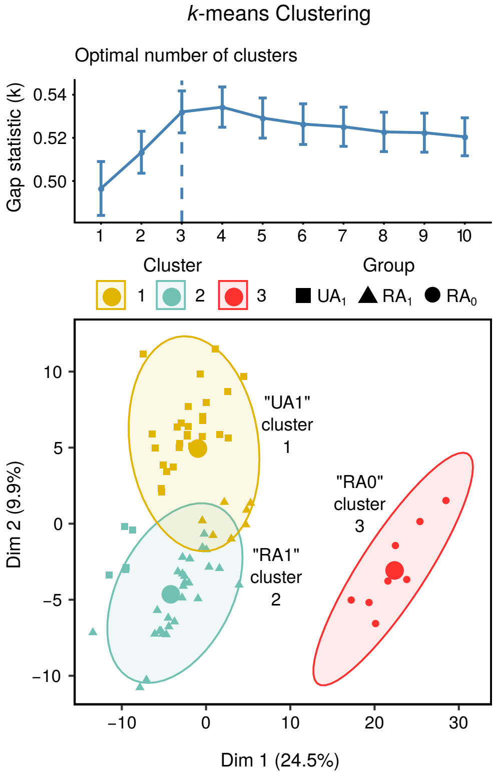


B

Sample assignment to clusters

| Cluster | Group | Cluster | Group | Cluster | Group |
|---|---|---|---|---|---|
| 1 | UA1 | 1 | UA1 | 2 | RA1 |
| 1 | UA1 | 1 | UA1 | 2 | RA1 |
| 1 | UA1 | 1 | UA1 | 2 | RA1 |
| 1 | UA1 | 1 | UA1 | 2 | RA1 |
| 1 | UA1 | 1 | UA1 | 2 | RA1 |
| 1 | RA1 | 1 | UA1 | 2 | RA1 |
| 1 | RA1 | 1 | UA1 | 2 | RA1 |
| 1 | RA1 | 1 | UA1 | 2 | RA1 |
| 1 | RA1 | 1 | RA1 | 2 | RA1 |
| 1 | RA1 | 2 | UA1 | 2 | RA1 |
| 1 | RA1 | 2 | RA1 | 2 | UA1 |
| 1 | UA1 | 2 | RA1 | 2 | UA1 |
| 1 | UA1 | 2 | RA1 | 2 | UA1 |
| 1 | UA1 | 2 | RA1 | 2 | UA1 |
| 1 | UA1 | 2 | RA1 | 2 | RA1 |
| 1 | UA1 | 2 | RA1 | 2 | RA1 |
| 1 | UA1 | 2 | RA1 | 2 | RA1 |
| 1 | UA1 | 2 | RA1 | 3 | RA0 |
| 1 | UA1 | 2 | RA1 | 3 | RA0 |
| 1 | UA1 | 2 | RA1 | 3 | RA0 |
| 1 | UA1 | 2 | RA1 | 3 | RA0 |
| 1 | UA1 | 2 | RA1 | 3 | RA0 |
| 1 | UA1 | 2 | RA1 | 3 | RA0 |
| 1 | UA1 | 2 | RA1 | 3 | RA0 |
| 1 | UA1 | 2 | RA1 | 3 | RA0 |
