## Supplementary material for "The DNA methylation Profile of Undifferentiated Arthritis Patients Anticipates their Subsequent Differentiation to Rheumatoid Arthritis": Supp. Figure Legends

**SUPPLEMENTARY FIGURE LEGENDS**

**Supplementary Figure 1.** Principal component analysis of top 100 most significant DMPs ($p \leq 2.8e-05$) between Group 1 (UA in 1987, UA in 2010) and Group 3 (RA in 1987, RA in 2010). PC1 and PC2 are shown, with 5.76% and 37.59% of variance explained, respectively. Coloring is set by group (Groups 1 to 4). Ellipsis show 95% confidence interval of the distribution of every group (minimum observations to plot an ellipsis = 3). Microarray model (450k or EPIC) was included as a covariate in the *limma* model.

**Supplementary Figure 2.** (A) Bar plots showing cell type composition proportions in HD and $UA_0$ groups. P values are derived from Mann-Whitney *U* mean comparison tests for each cell type and adjusted by FDR. (B) Examples of two selected DMRs. DMP, differentially methylated position; DVP, differentially variable position. (C) Venn diagram showing overlap of DMPs and DVPs in HD vs. $UA_0$ comparison.

**Supplementary Figure 3.** (A) Bar plots showing cell type composition proportions in $UA_1$ and $RA_1$ groups. P values are derived from Mann-Whitney *U* mean comparison tests for each cell type and adjusted by FDR. (B) Significantly enriched motifs in DMPs from both clusters, analyzed with HOMER. (C) Chromatin functional state enrichment based on public PBMC data from Roadmap Epigenomics Project. FDR, false discovery rate; TF, transcription factor; bHLH, basic helix-loop-helix domain family; CCAAT, CCAAT enhancer-binding protein family; NRF, nuclear respiratory factor family; Zf, zinc finger domain family.

**Supplementary Figure 4.** (A) ROC curves of SVM models with 0, 10 and 25 CpGs, with covariates RF and DAS44. (B) Accuracy of the prediction on the validation cohort across sequential numbers of CpGs as explanatory variables (1 to 50). ROC, receiver operating characteristics; AUC, area under the ROC curve.

**Supplementary Figure 5.** (A) Top, optimal number of *k* clusters determined by Gap statistic calculation. Bottom, PCA plot with the 3 clusters identified by *k*-means unsupervised clustering. Different clusters are highlighted in different colors, and the sample group is highlighted in different dot shapes. Ellipsis show 95% confidence interval of the distribution of every cluster. (B) Assignment of samples to each cluster. Clusters are colored with the same color as the most frequent group.
