## Supplementary material for "The DNA methylation Profile of Undifferentiated Arthritis Patients Anticipates their Subsequent Differentiation to Rheumatoid Arthritis": Supp. Table 1

**Supplementary Table 1.** Descriptive and clinical data of the patients included in this study.

| Sample_ID | Age_incl | Sex | sympt_dur_days | weight | length | TJC_DAS44 | SJC_DAS44 | ESR | CRP | RF | CCP2 | vaspain | vasda_patient | vaswb | DAS44 | DIAG_0_1987 | DIAG_1_1987 | DIAG_0_2010 | platform | ML_dataset |
|---|---|---|---|---|---|---|---|---|---|---|---|---|---|---|---|---|---|---|---|---|
| RA_1 | In 20s | F | 252 | 64.5 | 174 | 10 | 12 | 42 | 47 | Positive | Positive | 70 | 78 | 57 | 4.13 | RA | . | RA | EPIC | . |
| RA_2 | In 30s | F | 151 | 69.1 | 174 | 8 | 1 | 8 | 17 | Negative | Negative | . | . | . | . | RA | . | UA | EPIC | . |
| RA_3 | In 30s | F | 77 | 60 | 169 | 6 | 6 | 11 | 4 | Negative | Negative | 59 | 69 | 70 | 3.01 | RA | . | RA | EPIC | . |
| RA_4 | In 60s | F | 260 | 57 | 171 | 7 | 6 | 95 | 94 | Positive | Negative | 49 | 78 | 75 | 3.86 | RA | . | RA | EPIC | . |
| RA_5 | In 40s | M | 35 | 71 | 179 | 7 | 7 | 30 | 27 | Positive | Positive | 70 | 62 | 64 | 3.46 | RA | . | RA | EPIC | . |
| RA_6 | In 40s | M | 185 | 110 | 178 | 7 | 22 | 67 | 29 | Positive | Positive | 78 | 76 | 10 | 4.31 | RA | . | RA | EPIC | . |
| RA_7 | In 70s | M | 195 | 78 | 175 | 8 | 2 | 36 | 41 | Negative | Negative | 73 | 75 | 71 | 3.35 | RA | . | UA | EPIC | . |
| RA_8 | In 50s | F | 70 | 71 | 160 | 7 | 2 | 58 | 24 | Positive | Positive | 82 | 82 | 5 | 2.93 | RA | . | RA | EPIC | . |
| RA_9 | In 50s | F | 743 | 74 | 168 | 8 | 10 | 9 | 3 | Negative | Negative | 59 | 88 | 62 | 3.34 | RA | . | RA | EPIC | . |
| RA_10 | In 40s | F | 29 | 55 | 155 | 6 | 3 | 9 | 3 | Positive | Positive | 79 | 83 | 73 |  | RA | . | RA | EPIC | . |
| UA_1 | In 60s | M | 195 | 78 | 172 | 12 | 4 | 12 | 3 | Negative | Negative | 7 | 12 | 6 | 2.99 | UA | UA | RA | EPIC | validation |
| UA_2 | In 60s | F | . | 67 | 176 | 3 | 4 | 34 | 10 | Positive | Positive | 29 | 38 | 28 | 2.56 | UA | UA | RA | EPIC | validation |
| UA_3 | In 50s | M | 42 | 75 | 171 | 3 | 2 | 39 | 40 | Negative | Negative | 76 | 73 | 62 | 3.86 | UA | RA | UA | EPIC | validation |
| UA_4 | In 70s | M | . | 74.5 | 170 | 1 | 3 | 16 | 3 | Negative | Negative | 37 | 42 | 0 | 1.65 | UA | UA | UA | EPIC | validation |
| UA_5 | In 40s | F | 30 | 85 | 169 | 3 | 2 | 2 | 11 | Positive | Positive | 48 | 56 | 55 | 1.69 | UA | RA | RA | EPIC | validation |
| UA_6 | In 70s | F | 168 | 86 | 166 | 12 | 15 | 41 | 6 | Negative | Negative | 70 | 67 | 54 | 4.45 | UA | RA | RA | EPIC | validation |
| UA_7 | In 30s | M | 23 | . | . | 1 | 1 | 2 | 4 | Negative | Negative | 30 | 30 | 0 | 0.83 | UA | UA | UA | EPIC | validation |
| UA_8 | In 30s | M | 500 | 102 | 192 | 4 | 11 | 8 | 24 | Positive | Positive | 7 | 12 | 60 | 2.91 | UA | RA | RA | EPIC | validation |
| UA_9 | In 60s | F | 214 | 63 | 160 | 2 | 2 | 46 | 7 | Negative | Positive | 7 | 21 | 10 | 2.23 | UA | UA | RA | 450k | discovery |
| UA_10 | In 50s | F | 197 | 77.5 | 171 | 6 | 12 | 24 | 6 | Positive | Positive | 53 | 47 | 30 | 3.36 | UA | RA | RA | 450k | discovery |
| UA_11 | In 50s | F | 290 | 60.4 | 160 | 13 | 12 | 29 | 6 | Positive | Positive | . | . | . | . | UA | RA | RA | 450k | discovery |
| UA_12 | In 60s | M | 163 | 87 | 169 | 8 | 3 | 30 | 9 | Negative | Negative | 28 | 23 | 54 | 3.23 | UA | RA | UA | 450k | discovery |
| UA_13 | In 50s | M | 62 | 88 | 186 | 1 | 1 | 16 | 5 | Negative | Negative | 6 | 5 | 6 | 1.56 | UA | UA | UA | 450k | discovery |
| UA_14 | In 10s | F | 60 | 66 | 176 | 18 | 8 | 11 | 3 | Negative | Negative | 69 | 83 | 60 | 4.03 | UA | UA | RA | 450k | discovery |
| UA_15 | In 50s | M | 152 | 80 | 177 | 9 | 1 | 27 | 25 | Negative | Negative | 43 | 9 | 52 | 3.15 | UA | UA | UA | 450k | discovery |
| UA_16 | In 40s | F | 113 | 74 | 172 | 8 | 6 | 59 | 8 | Positive | Negative | 62 | 64 | 64 | 3.72 | UA | RA | RA | 450k | discovery |
| UA_17 | In 30s | F | 233 | 52 | 161 | 7 | 6 | 11 | 3 | Positive | Positive | 24 | 52 | 48 | 2.95 | UA | RA | RA | 450k | discovery |
| UA_18 | In 60s | M | 61 | 79.8 | 183 | 7 | 7 | 41 | 29 | Negative | Negative | 53 | 70 | 7 | 3.16 | UA | RA | RA | 450k | discovery |
| UA_19 | In 60s | F | 172 | 95 | 172 | 0 | . | 85 | 126 | Positive | Positive | 64 | 76 | 85 | . | UA | RA | RA | 450k | discovery |
| UA_20 | In 30s | F | 60 | 90 | 175 | 3 | 2 | 19 | 8 | Negative | Negative | 57 | 37 | 45 | 2.36 | UA | UA | UA | 450k | discovery |
| UA_21 | In 60s | M | . | 78.5 | 175 | 8 | 2 | 5 | 3 | Negative | Negative | 5 | 6 | 16 | 2.3 | UA | RA | RA | 450k | discovery |
| UA_22 | In 50s | M | 192 | 99 | 179 | 7 | 3 | 5 | 3 | Positive | Negative | 52 | 76 | 68 | 2.64 | UA | UA | RA | 450k | discovery |
| UA_23 | In 70s | M | 136 | 120 | 184 | 6 | 14 | 46 | 18 | Negative | Negative | 73 | 75 | 19 | 3.63 | UA | RA | RA | 450k | discovery |
| UA_24 | In 70s | M | 58 | 69 | 169 | 6 | 14 | 46 | 25 | Negative | Negative | 78 | 86 | 63 | 3.94 | UA | RA | RA | 450k | discovery |
| UA_25 | In 10s | M | . | 60 | 188 | 2 | 2 | 5 | 8 | Negative | Negative | 9 | 9 | 16 | 1.54 | UA | UA | UA | 450k | discovery |
| UA_26 | In 20s | M | 706 | 80 | 190 | 2 | 5 | 9 | 3 | Negative | Negative | 26 | 47 | 50 | 2.17 | UA | UA | UA | 450k | discovery |
| UA_27 | In 60s | F | 132 | 82 | 168 | 11 | 5 | 63 | 56 | Positive | Negative | 71 | 99 | 22 | 3.64 | UA | RA | RA | 450k | discovery |
| UA_28 | In 20s | F | 60 | 73 | 180 | 5 | 4 | 9 | 3 | Positive | Negative | 45 | 47 | 58 | 2.61 | UA | RA | RA | 450k | discovery |
| UA_29 | In 60s | M | 155 | 91 | 173 | 9 | 19 | 19 | 12 | Positive | Negative | 70 | 78 | 90 | 4.47 | UA | RA | RA | 450k | discovery |
| UA_30 | In 20s | F | . | 69 | 178 | 13 | 15 | 38 | 42 | Positive | Positive | 80 | 82 | 78 | 4.68 | UA | RA | RA | 450k | discovery |
| UA_31 | In 70s | F | 37 | 81.5 | 164 | 4 | 6 | 22 | 10 | Positive | Positive | 81 | 60 | 0 | 2.49 | UA | RA | RA | 450k | discovery |
| UA_32 | In 70s | F | 156 | 90 | 172 | 12 | 25 | 58 | 9 | Negative | Negative | 52 | 60 | 22 | 4.98 | UA | UA | RA | 450k | discovery |
| UA_33 | In 50s | F | 124 | 64 | 164 | 6 | 2 | 2 | 3 | Positive | Positive | 4 | 8 | 5 | 1.72 | UA | UA | RA | 450k | discovery |
| UA_34 | In 70s | M | 190 | 63 | 170 | 13 | 18 | 68 | 93 | Negative | Negative | 58 | 88 | 35 | 4.75 | UA | RA | RA | 450k | discovery |
| UA_35 | In 40s | F | 162 | 101 | 176 | 8 | 0 | 2 | 3 | Negative | Negative | 72 | 77 | 81 | 2.34 | UA | UA | RA | 450k | discovery |
| UA_36 | In 60s | F | . | 73 | 154 | 9 | 10 | 11 | 5 | Negative | Negative | 56 | 82 | 47 | 3.4 | UA | RA | RA | 450k | discovery |
| UA_37 | In 40s | M | 144 | 79 | 176 | 4 | 2 | 11 | 4 | Positive | Positive | 0 | 0 | 1 | 2.01 | UA | RA | RA | 450k | discovery |
| UA_38 | In 40s | F | 172 | 62 | 170 | 14 | 13 | 31 | 3 | Positive | Positive | 36 | 35 | 42 | 4.3 | UA | RA | RA | 450k | discovery |
| UA_39 | In 70s | F | 6 | . | . | . | . | 104 | 346 | Positive | Negative | . | . | . | . | UA | UA | UA | 450k | discovery |
| UA_40 | In 70s | F | 128 | . | . | 3 | 3 | 67 | 23 | Positive | Negative | 70 | 70 | 0 | 2.52 | UA | RA | RA | 450k | discovery |
| UA_41 | In 60s | M | 83 | 100 | 179 | 1 | 1 | 2 | 4 | Negative | Negative | 22 | 28 | 0 | 0.83 | UA | UA | UA | 450k | discovery |
| UA_42 | In 50s | M | 49 | 93 | 185 | 16 | 14 | 29 | 10 | Positive | Positive | 33 | 46 | 52 | 4.55 | UA | RA | RA | 450k | discovery |

| Sample_ID | Age_incl | Sex | sympt_dur_days | weight | length | TJC_DAS44 | SJC_DAS44 | ESR | CRP | RF | CCP2 | vaspain | vasda_patient | vaswb | DAS44 | DIAG_0_1987 | DIAG_1_1987 | DIAG_0_2010 | platform | ML_dataset |
|---|---|---|---|---|---|---|---|---|---|---|---|---|---|---|---|---|---|---|---|---|
| UA_43 | In 70s | F | 73 | 79 | 179 | 13 | 9 | 14 | 6 | Negative | Negative | 26 | 43 | 28 | 3.6 | UA | UA | RA | 450k | discovery |
| UA_44 | In 10s | F | 152 | 73 | 159 | 33 | 12 | 6 | 3 | Negative | Negative | . | . | . | . | UA | RA | RA | 450k | discovery |
| UA_45 | In 60s | M | 69 | 102 | 174 | 10 | 15 | 6 | 10 | Negative | Negative | 76 | 80 | 77 | 3.82 | UA | UA | RA | 450k | discovery |
| UA_46 | In 30s | F | 228 | 56.2 | 163 | 8 | 1 | 17 | 3 | Positive | Positive |  | 49 | 36 | 2.79 | UA | RA | RA | 450k | discovery |
| UA_47 | In 40s | M | 392 | 98 | 192 | 4 | 1 | 2 | 6 | Positive | Positive | 2 | 9 | 4 | 1.4 | UA | UA | RA | 450k | discovery |
| UA_48 | In 60s | F | 114 | 62 | 161 | 6 | 10 | 99 | 18 | Positive | Positive | 43 | 41 | 82 | 4.08 | UA | RA | RA | 450k | discovery |
| UA_49 | In 40s | M | 512 | 95 | 187 | 10 | 6 | 11 | 13 | Positive | Positive | 75 | 90 | 40 | 3.17 | UA | RA | RA | 450k | discovery |
| UA_50 | In 70s | M | 137 | 78 | 176 | 7 | 7 | 38 | 74 | Negative | Negative | 45 | 72 | 60 | 3.51 | UA | UA | UA | 450k | discovery |
| UA_51 | In 10s | M | 174 | 85 | 180 | 7 | 4 | 2 | 3 | Negative | Negative |  | 43 | 24 | 2.09 | UA | UA | RA | 450k | discovery |
| UA_52 | In 60s | M | 64 | 86 | 172 | 7 | 8 | 132 | 134 | Negative | Negative | 72 | 83 | 71 | 4.07 | UA | UA | RA | 450k | discovery |
| UA_53 | In 30s | M | 93 | 79 | 186 | 3 | 2 | 9 | 7 | Negative | Negative | 73 | 52 | 25 | 1.97 | UA | UA | UA | 450k | discovery |
| UA_54 | In 50s | M | 62 | 88 | 187 | 13 | 5 | 11 | 13 | Negative | Negative | 33 | 35 | 32 | 3.29 | UA | UA | RA | 450k | discovery |
| UA_55 | In 60s | M | 9 | 87 | 177 | 1 | 10 | 92 | 27 | Negative | Positive | 10 | 2 | 12 | 2.76 | UA | RA | RA | 450k | discovery |
| UA_56 | In 50s | F | 96 | 69 | 161 | 2 | 1 | 19 | 3 | Positive | Positive | 5 | 1 | 48 | 2.15 | UA | UA | RA | 450k | discovery |
| UA_57 | In 60s | F | 8 | 50 | 165 | 3 | 3 | 9 | 6 | Negative | Negative | 18 | 16 | 13 | 1.95 | UA | UA | UA | 450k | discovery |
| UA_58 | In 40s | F | 261 | 66 | 168 | 4 | 6 | 46 | 14 | Negative | Negative | 84 | 80 | 74 | 3.26 | UA | RA | UA | 450k | discovery |
| UA_59 | In 60s | M | 235 | . | . | 1 | 2 | 6 | 8 | Negative | Negative | 58 | 13 | 57 | 1.67 | UA | RA | UA | 450k | discovery |
| UA_60 | In 40s | F | 274 | 72 | 172 | 8 | 4 | 6 | 3 | Negative | Negative | 55 | 71 | 2 | 2.39 | UA | UA | UA | 450k | discovery |
| UA_61 | In 30s | F | 182 | 102 | 176 | 0 | 0 | 14 | 18 | Negative | Positive | 0 | 0 | 0 | 0.87 | UA | UA | RA | 450k | discovery |
| UA_62 | In 40s | F | 119 | 71 | 170 | 2 | 1 | 24 | 21 | Positive | Positive | 74 | 64 | 68 | 2.37 | UA | RA | RA | 450k | discovery |
| UA_63 | In 40s | F | 38 | 69 | 168 | 6 | 1 | 33 | 21 | Negative | Positive | 34 | 25 | 41 | 2.84 | UA | UA | RA | 450k | discovery |
| UA_64 | In 50s | F | 247 | 85 | 176 | 7 | 5 | 9 | 4 | Negative | Negative | 27 | 34 | 14 | 2.58 | UA | RA | RA | 450k | discovery |
| UA_65 | In 80s | F | 158 | 68 | 163 | 0 | 6 | 39 | 3 | Negative | Negative | 54 | 76 | 9 | 1.66 | UA | RA | UA | 450k | discovery |
| UA_66 | In 50s | F | 318 | . | 160 | 15 | 11 | 50 | 49 | Positive | Positive | 72 | 85 | 24 | 4.26 | UA | RA | RA | 450k | discovery |
| UA_67 | In 60s | M | 77 | 93 | 176 | 10 | 11 | 53 | 58 | Negative | Negative | 44 | 57 | 4 | 3.76 | UA | RA | RA | 450k | discovery |
| UA_68 | In 70s | F | 161 | 61 | 163 | 11 | 14 | 9 | 8 | Negative | Negative | 72 | 70 | 44 | 3.74 | UA | UA | RA | 450k | discovery |
| UA_69 | In 40s | F | . | . | . | 2 | 2 | 2 | 3 | Negative | Negative | 75 | 80 | 96 | 1.81 | UA | UA | UA | 450k | discovery |
| UA_70 | In 60s | F | 253 | . | . | 12 | 5 | 22 | 18 | Negative | Positive | 60 | 70 | 50 | 3.57 | UA | RA | RA | 450k | discovery |
| UA_71 | In 60s | F | 48 | 79 | 173 | 2 | 0 | 11 | 3 | Negative | Negative | 49 | 49 | 25 | 1.73 | UA | UA | RA | 450k | discovery |
| UA_72 | In 70s | M | 110 | 82 | 181 | 5 | 10 | 58 | 119 | Positive | Negative | . | . | . | . | UA | RA | RA | 450k | discovery |
| HD_1 | In 20s | F | . | . | . | . | . | . | . | . | . | . | . | . | . | HD | . | . | 450k | . |
| HD_2 | In 20s | M | . | . | . | . | . | . | . | . | . | . | . | . | . | HD | . | . | 450k | . |
| HD_3 | In 30s | M | . | . | . | . | . | . | . | . | . | . | . | . | . | HD | . | . | 450k | . |
| HD_4 | In 40s | M | . | . | . | . | . | . | . | . | . | . | . | . | . | HD | . | . | 450k | . |
| HD_5 | In 30s | M | . | . | . | . | . | . | . | . | . | . | . | . | . | HD | . | . | 450k | . |
| HD_6 | In 30s | F | . | . | . | . | . | . | . | . | . | . | . | . | . | HD | . | . | 450k | . |
| HD_7 | In 40s | M | . | . | . | . | . | . | . | . | . | . | . | . | . | HD | . | . | 450k | . |
| HD_8 | In 40s | M | . | . | . | . | . | . | . | . | . | . | . | . | . | HD | . | . | 450k | . |
| HD_9 | In 20s | F | . | . | . | . | . | . | . | . | . | . | . | . | . | HD | . | . | 450k | . |
| HD_10 | In 20s | M | . | . | . | . | . | . | . | . | . | . | . | . | . | HD | . | . | 450k | . |
| HD_11 | In 20s | F | . | . | . | . | . | . | . | . | . | . | . | . | . | HD | . | . | 450k | . |
| HD_12 | In 20s | M | . | . | . | . | . | . | . | . | . | . | . | . | . | HD | . | . | 450k | . |
| HD_13 | In 20s | M | . | . | . | . | . | . | . | . | . | . | . | . | . | HD | . | . | 450k | . |

Age_incl, age at the time of inclusion in the study; sympt_dur_days, symptoms duration in days; TJC_DAS44, tender joint count; SJC_DAS44, swollen joint count; ESR, erythrocyte sedimentation rate; CRP, C-reactive protein; RF, seropositivity for rheumatoid factor (qualitative); CCP2, seropositivity for anti-cyclic citrullinated peptide antibody (qualitative); vaspain, pain (mm); vasda_patient, disease activity (mm) ; vaswb, wellbeing (mm); DAS44, disease activity score 44; DIAG_0_1987, diagnosis at time 0 by ACR 1987 criteria; DIAG_1_1987, diagnosis after 1 year of time 0 by ACR 1987 criteria; DIAG_0_2010, diagnosis at time 0 by ACR/EULAR 2010 criteria; platform, Illumina methylation array model; ML_dataset, Discovery or validation dataset for the machine learning (ML) predictive models.
